# Whole-genome sequencing of SARS-COV-2 showed wide spread of B.1.525 in February 2021 in Libya

**DOI:** 10.1101/2021.07.03.21257942

**Authors:** Inas M Alhudiri, Ahmad M. Ramadan, Khaled M Ibrahim, Adel Abdalla, Mouna Eljilani, Mohamed Ali Salem, Hajer Mohamed Elgheriani, Salah Edin El Meshri, Adam Elzagheid

## Abstract

Alpha (B.1.1.7) SARS-COV-2 variant was detected in September 2020 in minks and humans in Denmark and UK. This variant has several mutations in the spike region (S) which could increase the transmissibility of the virus 43-90% over previously circulating variants. The National Center for Disease Control (NCDC) announced on 24th February 2021 a 25% frequency of B.1.1.7 strain in Libya using a reverse-transcriptase quantitative PCR assay. This assay relies on the specific identification of the H69-V70 deletion in S gene which causes its failure of amplification (SGTF). This deletion is not specific for B.1.1.7; but is also characteristic of two other SARS-COV-2 variants. This study aimed to estimate the frequency of B.1.1.7 and identify other variants circulating in Libya in February 2021. We performed whole genome sequencing of 67 positive SARS-COV-2 samples collected on 25th February 2021 in Libya which were also tested by RT-qPCR for SGTF. Our results showed that 55% of samples had mutations specific to B.1.525 strain and only ∼3% of samples belonged to B.1.1.7. These findings suggested that B.1.525 was spreading widely in Libya. The use of such RT-qPCR assay although useful to track some variants, it cannot discriminate between variants with H69-V70 deletion. RT-qPCR assays could be multiplexed to identify multiple variants and screen samples prior to sequencing. We emphasize on the need for providing whole-genome sequencing to the main COVID-19 diagnostic laboratories in Libya as well as establishing international collaboration for building capacity and advancing research in this time of the pandemic.

---

A SARS-CoV-2 spike gene (S) deletion (H69-V70) was detected in cluster-5 variant in minks and humans in September 2020 (named lineage B.1.1.7).^1^ This variant has several mutations in the spike region (S) which could increase the transmissibility of the virus 43-90% over previously circulating variants.^2^ Whole genome sequencing is considered the gold method for identifying such variants. However, reverse-transcriptase quantitative PCR assays (RT-qPCR) targeting specific mutations in the S gene were considered useful surrogates for tracking B.1.1.7 in different studies ^3,4^. This study aimed to estimate the frequency of B.1.1.7 and identify other variants circulating in Libya in February 2021.

The national center for disease control (NCDC) announced on 24^th^ February 2021 the discovery of B.1.1.7 strain in Libya using a reverse-transcriptase PCR assay (SARS-COV-2 UK RT-PCR kit, Vircell) for S-gene target failure (SGTF) and reported that 25% of the tested samples were UK variant.^5^ This assay relies on the specific identification of the 69-70 deletion in S gene which causes S gene drop out in RT-PCR; characteristic of the UK variant (B.1.1.7).

We analyzed 67 SARS-COV-2 positive samples collected on 25^th^ February to track the presence of the UK variant using TaqPath™ COVID□19 CE□IVD RT□PCR Kit (Thermo Fisher Scientific). Median cycle threshold of these samples was 25 ± 4.6 cycles. To confirm the results, we sent the extracted RNA of these samples to a genomic analysis laboratory in Spain for whole-genome sequencing (WGS) because of limited sequencing facilities in Libya. Next generation sequencing of libraries was done using Illumina platform followed by sequence mapping using Sophia MMD Software. Determination of strains, number of variants in coding and non-coding regions, as well as evolutionary origin and geolocation was performed through Nextclade and Pangolin applications^6^.

No amplification of S gene (SGTF) was seen in 67% of samples (45/67). This result was initially interpreted as the frequency of the UK variant in Libya according to SGTF results. However, these results did not coincide with the epidemiological status of the disease which showed constant disease prevalence since December 2020 with no noticeable increase.^7^ Unexpectedly, our analysis showed that ∼ 3% (2/67) of samples were B.1.1.7 and slightly over half of samples were B.1.525 strain (37/67, 55%). The findings also demonstrated that within the samples with SGTF, 80% (36/45) had ΔH69/ΔV70 after confirmation by sequencing. Table 1 describes the different variants detected in our samples.

**Table 1.** SARS-COV-2 lineages identified in samples from Libyan patients

| Lineage | Frequency<br>n=67 (%) | Reason for<br>classification |
| --- | --- | --- |
| B.1.525 | 37 (55) | Pangolin Tool<br>assignment<sup>9</sup> |
| B.1.1.7 | 2 (2.9) | Pangolin Tool<br>assignment<sup>13</sup> |
| A.27 | 10 (14.9) | Pangolin Tool<br>assignment<sup>14</sup> |
| B.1 | 6 (8.9) | Pangolin Tool<br>assignment<sup>15</sup> |
| Other<br>Miscellaneous | 12 (17.9) |  |

B.1.525 lineage also contains E484K associated with South African variant and immune escape.^3^ This mutation was found in all B.1.525 strains and 62.7% (42/67) of total samples. World health organization designated B.1.525 as variant of interest on 17. Mar. 2021 and has been detected in multiple countries including Nigeria, UK, USA and few in Tunisia and Morocco.^8^ Other key mutations include ORF1a; del3675/3677 and S;F888L^9^. On the other hand, B.1.1.7 mutations detected in the S protein were deletions at residues 69/70 and 144, and other substitutions including N501Y and D614G.^10^ There are other defining mutations in ORF1a and N genes in addition to those in S gene^10^. The inclusion of mutations not common among variants could help differentiate between them in multiplex assays, e.g., an assay targeting deletion (H69-V70) and E484K could discriminate between B.1.525 and B.1.1.7 lineages.

Studies using RT-qPCR assays conducted in different countries have suggested SGTF as important surrogate marker for B.1.1.7 but several factors should be taken into consideration including the time and place of testing as well as community/hospital setting^3,11^.

As of the beginning of June 2021, our routine screening for SARS-CoV-2 variants using Taqpath assay interestingly revealed a noticeable increase in the frequency of the S-gene in positive samples. The S-gene was detected in over 50% of positive samples (data from our lab, no published). Sequencing data from other studies showed that the detection of S-gene is a characteristic feature of the Delta Variant of Concern.^12^ Unfortunately, whole genome sequencing was not undertaken to confirm the results.

COVID-19 testing in Libya is performed solely by government laboratories. These laboratories lack whole genome sequencing facilities and experience. The SARS-COV-2 diagnostic laboratories across Libya currently test approximately 6000 samples per day using RT-qPCR assay.

These findings suggested that B.1.525 was spreading widely in Libya. The use of such RT-qPCR assay although useful to track some variants, it cannot discriminate between variants with H69-V70 deletion. RT-qPCR assays could be multiplexed to identify multiple variants and screen samples prior to sequencing. We emphasize on the need for providing whole-genome sequencing to the main COVID-19 diagnostic laboratories in Libya as well as establishing international collaboration for building capacity and advancing research in this time of the pandemic.

## Data Availability

SARS-COV-2 sequencing data are publicly available via the link provided.

https://www.ncbi.nlm.nih.gov/nuccore/?term=BTRC+Libya

## Acknowledgment

The authors gratefully acknowledge the support of ministry of health, ministry of higher education, the Libyan Embassy in Spain and the Libyan authority for research, science and technology and all lab researchers at Biotechnology Research Center COVID-19 detection team.

## Author contributions

IA and AR are joint first authors who conceptualized the project and wrote the original draft. KI, S E and ME reviewed and edited the draft. AA, MS and HE collected, analyzed and interpreted RT-PCR data. AR, IA and KI analyzed and interpreted whole genome sequencing data. AE acquired the financial support for this project and reviewed the draft.

## Ethical approval

Ethical approval All procedures performed in studies involving human participants were in accordance with the ethical standards of the institutional and/or national research committee and with the 1964 Helsinki declaration and its later amendments or comparable ethical standards. This study was approved by the Biotechnology Research Center Bioethics Committee.

